# Trends and opportunities in tick-borne disease geography

**DOI:** 10.1101/2021.01.28.21250676

**Authors:** Catherine A. Lippi, Sadie J. Ryan, Alexis L. White, Holly D. Gaff, Colin J. Carlson

## Abstract

Tick-borne diseases are a growing problem in many parts of the world, and their surveillance and control touches on challenging issues in medical entomology, agricultural health, veterinary medicine, and biosecurity. Spatial approaches can be used to synthesize the data generated by integrative One Health surveillance systems, and help stakeholders, managers, and medical geographers understand the current and future distribution of risk. Here, we performed a systematic review of over 8,000 studies, and identified a total of 303 scientific publications that map tick-borne diseases using data on vectors, pathogens, and hosts (including wildlife, livestock, and human cases). We find that the field is growing rapidly, with the major *Ixodes-*borne diseases (Lyme disease and tick-borne encephalitis in particular) giving way to monitoring efforts that encompass a broader range of threats. We find a tremendous diversity of methods used to map tick-borne disease, but also find major gaps: data on the enzootic cycle of tick-borne pathogens is severely underutilized, and mapping efforts are mostly limited to Europe and North America. We suggest that future work can readily apply available methods to track the distributions of tick-borne diseases in Africa and Asia, following a One Health approach that combines medical and veterinary surveillance for maximum impact.

## Introduction

Tick-borne diseases are increasingly recognized as a neglected subset of emerging infections. The expansion of Lyme disease (Lyme borreliosis) in the United States and Europe has brought attention to the ecological dimensions of their emergence, and the broader links between global change and the expansion and resurgence of vector-borne disease. More recently, in the United States, the spread of Powassan virus and spotted fever group rickettsioses have been seen as evidence of an emerging trend: tick-borne pathogens are proliferating, spreading to new areas, and emerging in human populations, at a comparable rate to other zoonotic threats (Woolhouse et al. 2008, Smith et al. 2014). These perspectives, of course, focus predominantly on Western countries, where zoonotic diseases have a comparatively lower burden (Torgerson and Macpherson 2011, Kuris 2012).

Worldwide, tick-borne diseases are a persistent example of problems at the One Health interface between humans, wildlife, and agriculture. Many, like Crimean-Congo haemorrhagic fever and tick-borne encephalitis, are a particularly significant problem for impoverished livestock keepers in rural locations (Grace et al. 2017, Espinaze et al. 2018). These conditions are severely neglected, often receiving less clinical attention and dedicated public health funding than directly-transmitted zoonotic viruses like influenza (Tick-Borne Disease Working Group 2018). Ticks are usually prioritized below mosquitoes by vector control programs, given the lower comparative public health burden and often limited agency resources. Vector control to reduce tick populations is further limited by the availability of large-scale mitigation strategies and control technologies, where additional research may be required to develop effective control measures for ticks and wildlife hosts (White and Gaff 2018, Rochlin et al. 2019, Eisen and Stafford 2020). Prevention and treatment in clinical settings is similarly limited: despite their frequently severe prognosis and high case fatality rate, few tick-borne pathogens have available or widely used vaccines, and only one research laboratory in the world regularly works with tick-borne pathogens at BSL4 containment. Detection and diagnosis of many tick-borne infections in humans is also challenging, given the broad clinical presentation of many tick-borne diseases, current availability of reliable diagnostic tests, and multi-tiered approaches needed to confirm pathogens (Fatmi et al. 2017, Bush and Vazquez-Pertejo 2018).

The relative neglect of tick-borne illnesses among vector-borne diseases is also evident in basic disease surveillance data. Lyme surveillance is well established, and several large clinical datasets have been curated and used by researchers; and a handful of tick-borne zoonoses are notifiable in systems like ProMed-mail and the WHO Disease Outbreak News. But on the whole, tick-borne pathogens are severely undersurveilled, meaning many outbreaks likely go unreported, and the distributions and burdens of these diseases are likely underestimated or entirely unknown. In no small part this reflects the unique challenges of acquiring and verifying data on tick-borne diseases for spatial analyses. The vectors and within-vector pathogens may have distinct distributions, and human case data may also differ in its distribution, as a function of encounter and exposure. In addition, vector and human case data require different methods of data collection, each with logistical constraints that may dictate the geographic extent of sampling. Given these challenges, tick distribution maps are often used as a proxy for either transmission exposure risk, or to describe the human disease distribution, but vector range is poorly characterized for most tick-borne pathogens. Moreover, all tick-borne infections of humans are zoonotic, and many have wildlife hosts, where data on infection in each layer of human, domesticated, and wildlife host, describe different components of the transmission process. Data on animal reservoirs are not always collected by existing health surveillance networks, and are rarely stored in the same geoinformatic systems.

Maps are a primary tool for visualizing spatial information regarding pathogens and communicating potential risk of exposure. Disease maps have long been used in public health to describe the distribution of vector-borne diseases, ranging in complexity from plotted cases (i.e. dot maps) to projected risk predictions modeled with machine learning algorithms. Regardless of complexity, mapped products rely on the availability of georeferenced datasets. Given the challenges surrounding tick-borne disease research, we hypothesized that most tick-borne diseases have not been comprehensively mapped. To evaluate the state of the field, we performed a systematic literature review and identified all studies of tick-borne pathogens that produced spatial data, models, or other mapping analyses of the pathogens themselves, or used maps of the vectors as a proxy. We found that, despite the obvious threat posed to human and animal health by these diseases - and their growing significance in a changing world - the vast majority are undermapped, and many pathogens have not been mapped at all. Based on our results, we identify trends in the field, including shifting priorities for surveillance and methodological innovation, and discuss where surveillance efforts may need to be supplemented in the coming years.

## Methods

We compiled a list of twenty-seven tick-borne pathogens for inclusion in literature searches, using data from (Dantas-Torres et al. 2012) and (Brackney and Armstrong 2016). Four additional pathogens of recent public health interest were also included for review: *Borrelia mayonii, B. miyamotoi, Rickettsia parkeri* (CDC 2018), and Panola mountain ehrlichiosis (*Ehrlichia* spp.) (Reeves et al. 2008). A final list of pathogens and vectors included in the study is available in Tables 1 and 2.

**Table 1.** Viruses included in the study.

| Pathogen | Family | Vectors |
| --- | --- | --- |
| African Swine Fever Virus | Asfarviridae | *Ornithodoros* spp. |
| Bourbon Virus | Orthomyxoviridae | *Amblyomma* spp. |
| Colorado Tick Fever Virus | Reoviridae | N/A |
| Crimean-Congo Hemorrhagic Fever Virus | Bunyaviridae | *Hyalomma* spp.<br>*Rhipicephalus* spp. |
| Heartland Virus | Bunyaviridae | *Amblyomma* spp.<br>*Haemaphysalis* spp. |
| Huaiyangshan Banyangvirus | Bunyaviridae | *Haemaphysalis* spp. |
| Kyasanur Forest Disease Virus | Flaviviridae | *Haemaphysalis* spp. |
| Louping Ill Virus | Flaviviridae | *Ixodes* spp. |
| Nairobi Sheep Disease Virus | Bunyaviridae | *Haemaphysalis* spp. |
| Omsk Hemorrhagic Fever Virus | Flaviviridae | N/A |
| Powassan Virus | Flaviviridae | *Ixodes* spp. |
| Sawgrass Virus | Rhabdoviridae | N/A |
| Tick-Borne Encephalitis Virus | Flaviviridae | *Dermacentor* spp.<br>*Ixodes* spp. |

**Table 2.**
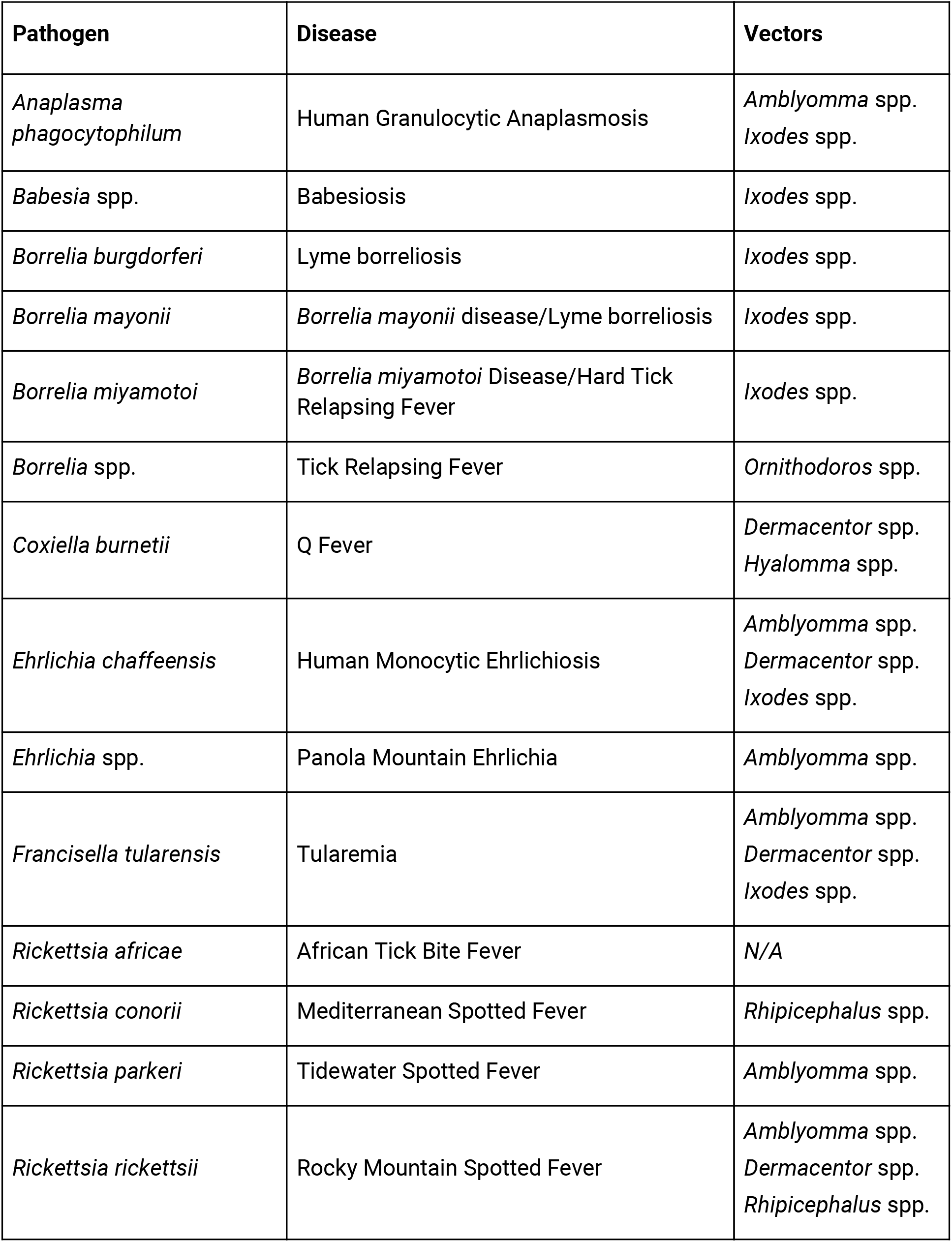
Bacteria and protozoan parasites included in the study.

We conducted literature searches following Preferred Reporting Items for Systematic Reviews and Meta-analyses (PRISMA) statement guidelines, a checklist of criteria to ensure transparency in systematic reviews (Liberati et al. 2009, Moher 2009). Searches for each pathogen, and named diseases they cause, were conducted in the PubMed Central (PMC) and Google Scholar databases from January-September 2020. The search queries used included combinations of pathogen names and key terms used to describe mapping and spatial analysis studies, taking the format: [species name] OR [disease name] AND (“SaTScan” OR “MaxEnt” OR “spatial cluster^*^”OR “spatial analysis” OR “geospatial” or “ecological niche model^*^” OR “mapping” OR “nearest neighbor” OR “spatial GLM^*^” OR “species distribution model^*^”). We did not place restrictions on geographic region of study or date of publication, and searches were limited to English language results. Additional novel records for screening were taken from cited literature in records identified via database searches.

Duplicate records were removed from search results, and the remaining papers were screened for further review. Original, peer-reviewed studies with a spatial component, and within the taxonomic scope of the review, were assessed for further screening. The remaining full-text studies were reviewed for inclusion. Literature reviews, expert commentaries, synthesis papers, conference abstracts, and unpublished theses were excluded from results, as were studies using serology not resolved to the taxonomic level of interest, and studies that focused on pathogens solely known for their veterinary importance. We recorded the citation, DOI link, geographic region, pathogens, vectors, data sources and sampling methods for vectors, and data inputs for each study included in our final dataset.

Mapping and spatial analysis methods were also recorded for papers. In order to describe the types of maps in the studies, we created a key, based on a previous study of helminth parasite mapping (Schluth et al. 2021), and classified studies into eight types. Studies could contain more than one type of map (Table 3).

**Table 3.** Eight types of study methodologies defined in this review.

| Type of Study | Definition (example) |
| --- | --- |
| Cluster Analysis | Any type of cluster analysis was used, including SatScan cluster analysis, kernel density hotspot modeling, or similar, e.g.(15) |
| Ecological Niche Modeling | A species distribution modeling (SDM) algorithm was applied to point data of occurrences of ticks or tick borne disease, and the resulting map was a function of environmental drivers of geographic distributions |
| Endemicity Mapping | Mapping the extent of ticks or tick-borne disease occurrence, based on a systematic or manual review of historical or published data and expert opinion, typically expressed with administrative boundaries or zones of suspected risk |
| Genetic Mapping | Maps which included locations of phylogenetic descriptions - e.g. a pie chart of strain type frequency at a given location |
| Point Data | Spatial data points of information (e.g. incidence of human cases, presence or absence of vectors), presented on a map in a format accessible for reuse through digitization |
| Prevalence Mapping | Maps of tick-borne disease prevalence, in humans or other hosts, visualized using raw (unaltered and unmodeled) data |
| Prevalence Modeling | Maps generated as predicted functions of prevalence through some sort of quantitative modeling |
| Risk Mapping | Projection of a modeled output (such as linear regression model output) onto a continuous geographic area or region, intended to communicate the geographic extent and intensity of transmission risk |

### Limitations

Limitations to this study include the potential for gaps in coverage in certain geographic regions, reflecting limiting our searches to English-language publications. These gaps may be particularly evident in countries with long histories of vector-borne disease management, such as China and Russia, that have extensive bodies of research not readily accessible due to language barriers in the literature (Ruzek et al. 2019, Zhang et al. 2019). While we included many types of mapping approaches, and attempted to describe the range of those approaches, those that we perceived as not quantitative were excluded, as was grey literature such as reports and conference abstracts.

## Results

Our initial database searches returned 12,482 records, which yielded 8,608 unique publications. An overview of the literature screening process, following PRISMA guidelines, is shown in Fig. 1. The final screened dataset comprised 303 studies on tick-borne pathogens with a mapped spatial component. Four out of 27 pathogens of interest did not have any associated mapping studies that met our screening criteria: Omsk hemorrhagic fever virus, sawgrass virus, Colorado tick fever virus, and *Rickettsia africae*. Only nine pathogens had more than 10 associated mapping studies: ASF virus, CCHF virus, TBE virus, *Borrelia burgdorferi, Anaplasma phagocytophilum, Coxiella burnetii, Ehrlichia chaffeensis, Francisella tularensis*, and *Rickettsia rickettsii*. While the majority of studies focused on Lyme disease (40.26%) or tick-borne encephalitis (15.51%), the overall number of published work with a mapping component has increased dramatically across taxa in the past decade (Fig. 2). Tick vectors from seven genera were represented in the final dataset: *Amblyomma, Dermacentor, Haemaphysalis, Hyalomma, Ixodes, Ornithodoros, and Rhipicephalus* (Fig. 2). Studies with data from *Ixodes* were the most prevalent, featured in 65.42% of studies including information on the vector. These typically focused on three species of medical concern: *Ixodes scapularis* (Say, Ixodida: Ixodidae), *Ixodes ricinus* (L., Ixodida: Ixodida), *Ixodes pacificus* (Cooley & Kohls, Ixodida: Ixodidae).

**Figure 1.**
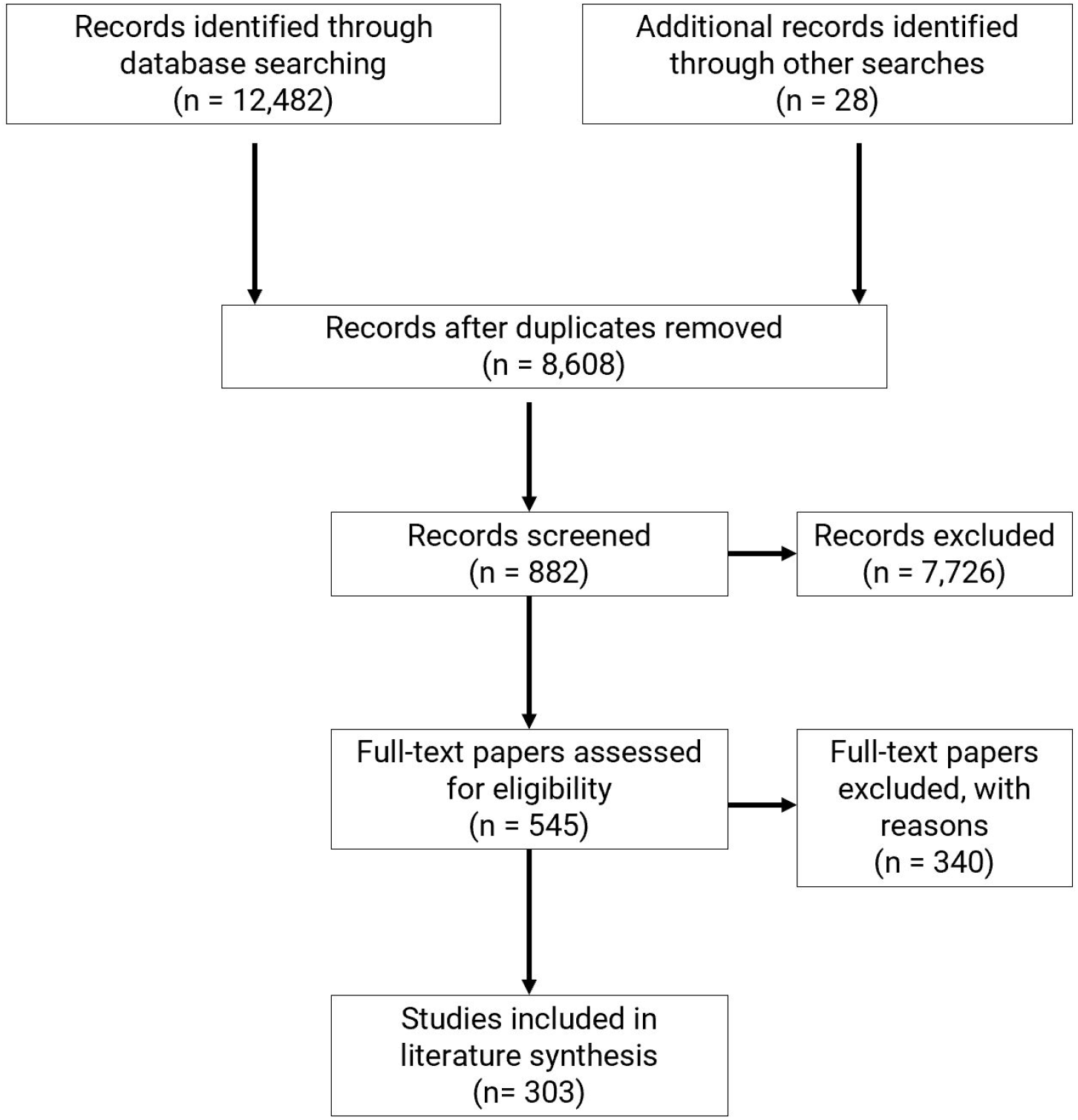
PRISMA flow diagram outlining the literature search and screening process.

**Figure 2.**
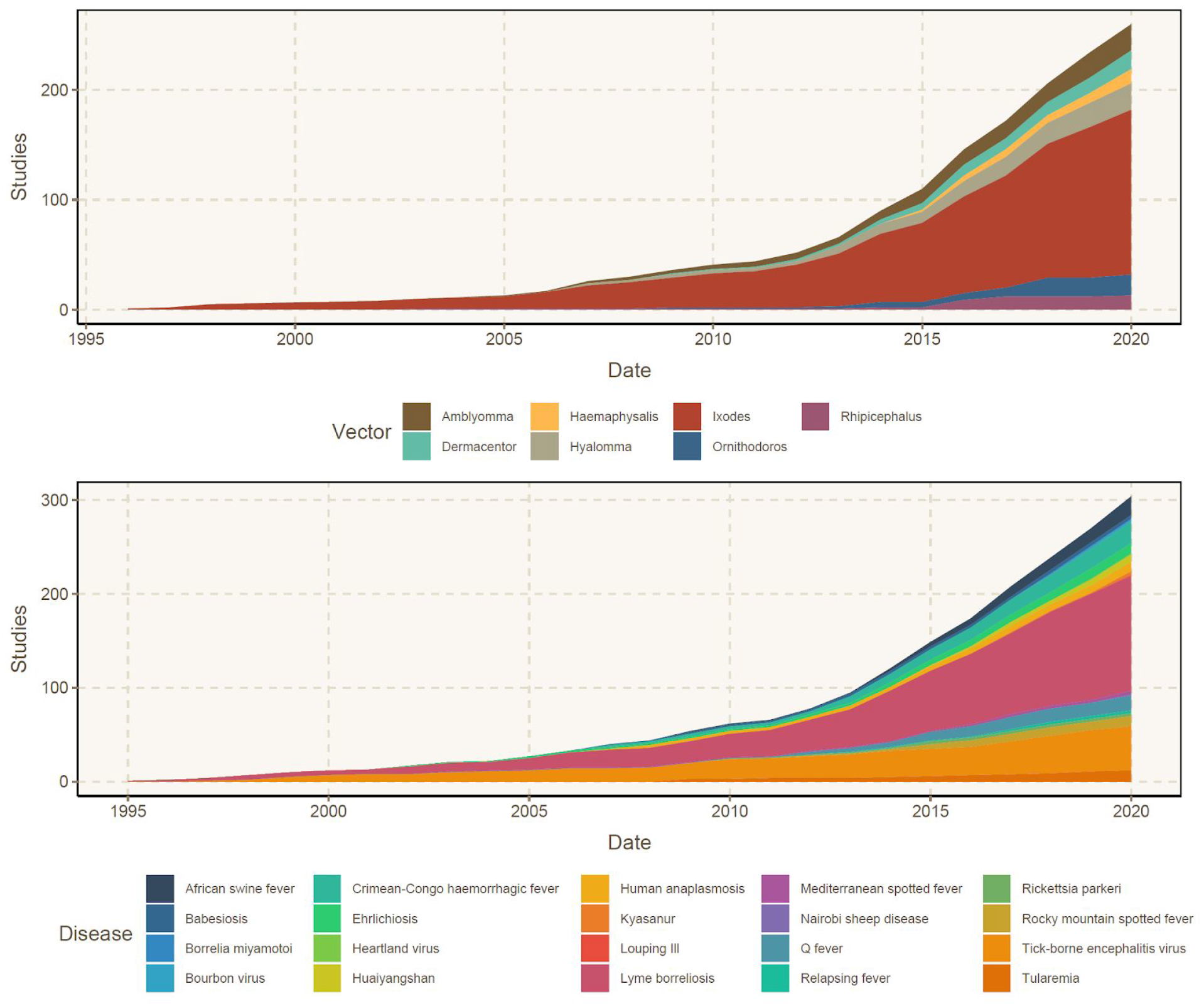
The cumulative number of studies that collected data about a given genus of tick vector (top) or tick-borne disease (bottom).

The eight mapping approaches (Table 3) used to classify studies were all represented in the final dataset of screened papers (Fig. 3). Mapping raw occurrence points of pathogens and vectors was the most frequently used approach in communicating spatial information, and was used in nearly half (47.85%) of screened studies. Risk mapping (31.02%) and endemicity mapping (29.70%) were also commonly used to communicate the spatial distribution of tick-borne pathogens or risk of exposure to ticks. Ecological niche modeling was used to estimate distributions, typically for tick vectors, in 22.11% of studies, and the majority (76.11%) of these studies produced niche models with the maximum entropy (MaxEnt) method (Phillips et al. 2006). Cluster analysis was used in 18.48% of studies, where tests for spatial autocorrelation (n=15) and spatial scanning statistics implemented in SaTScan (n=31) were frequently used for cluster detection. Prevalence modeling (8.58%) and genetic mapping (4.62%) were the methods least used in the final dataset of screened literature.

**Figure 3.**
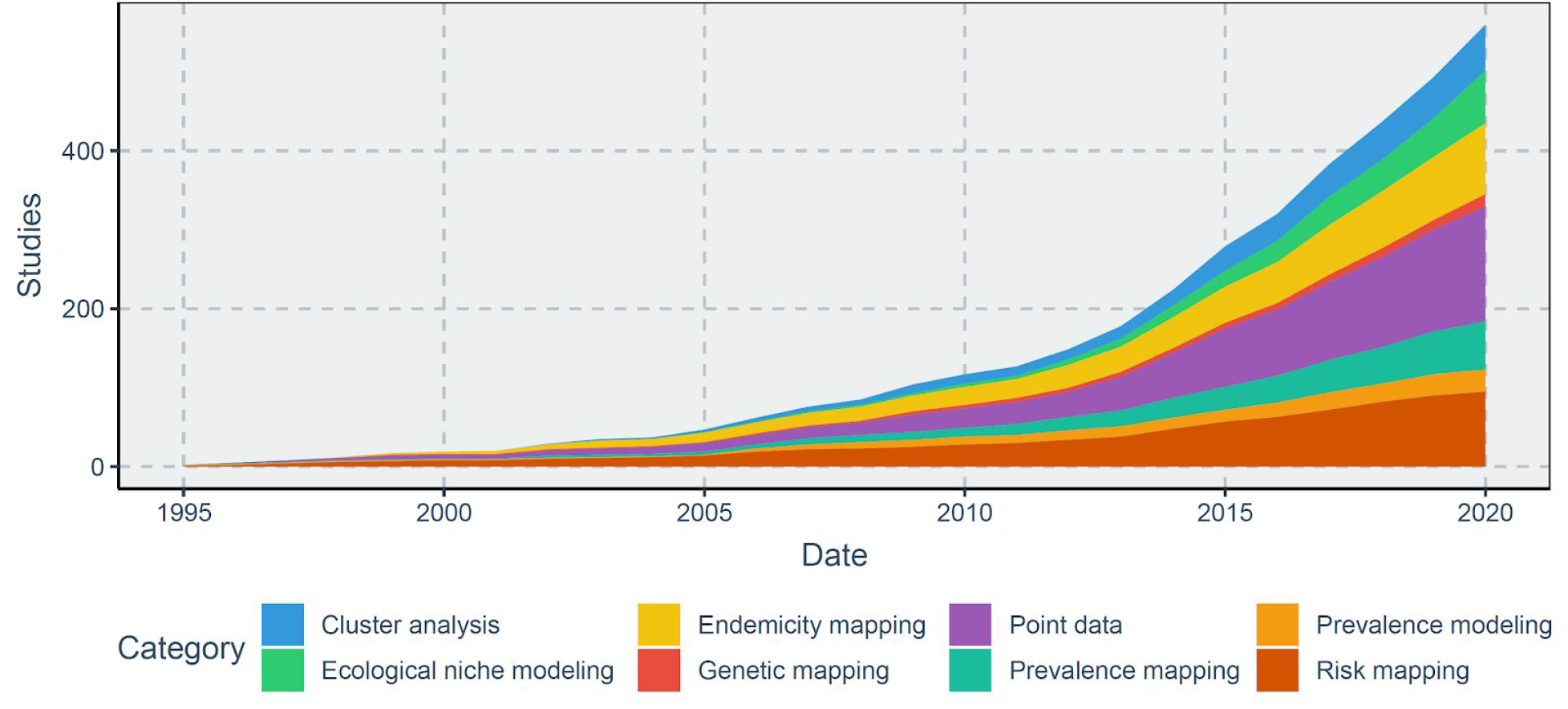
The cumulative number of studies using any of eight given methodologies.

Data sources used to generate maps varied between studies, where 74.92% used pathogen records as inputs, and a quarter (25.08%) used tick vectors as proxies for the pathogens they transmit (Fig. 4). Previously published datasets were used in 20.79% of studies, where resources including museum records, online databases, and literature reviews were commonly leveraged as data sources for spatial analyses. Human cases were used as data inputs in 39.60% of mapping efforts, and data from other vertebrate hosts such as domesticated livestock (11.88%) and wildlife (9.57%) were less common (Fig. 4). The relative proportions of domesticated and wildlife hosts sampled for pathogens across studies are presented in Fig. 5. Livestock sampled for pathogens in the literature largely consisted of hoofstock (i.e. cattle, sheep, and pigs) and domestic dogs; rodents, ungulates, and suids were the groups most frequently sampled for pathogens in wildlife serology studies.

**Figure 4.**
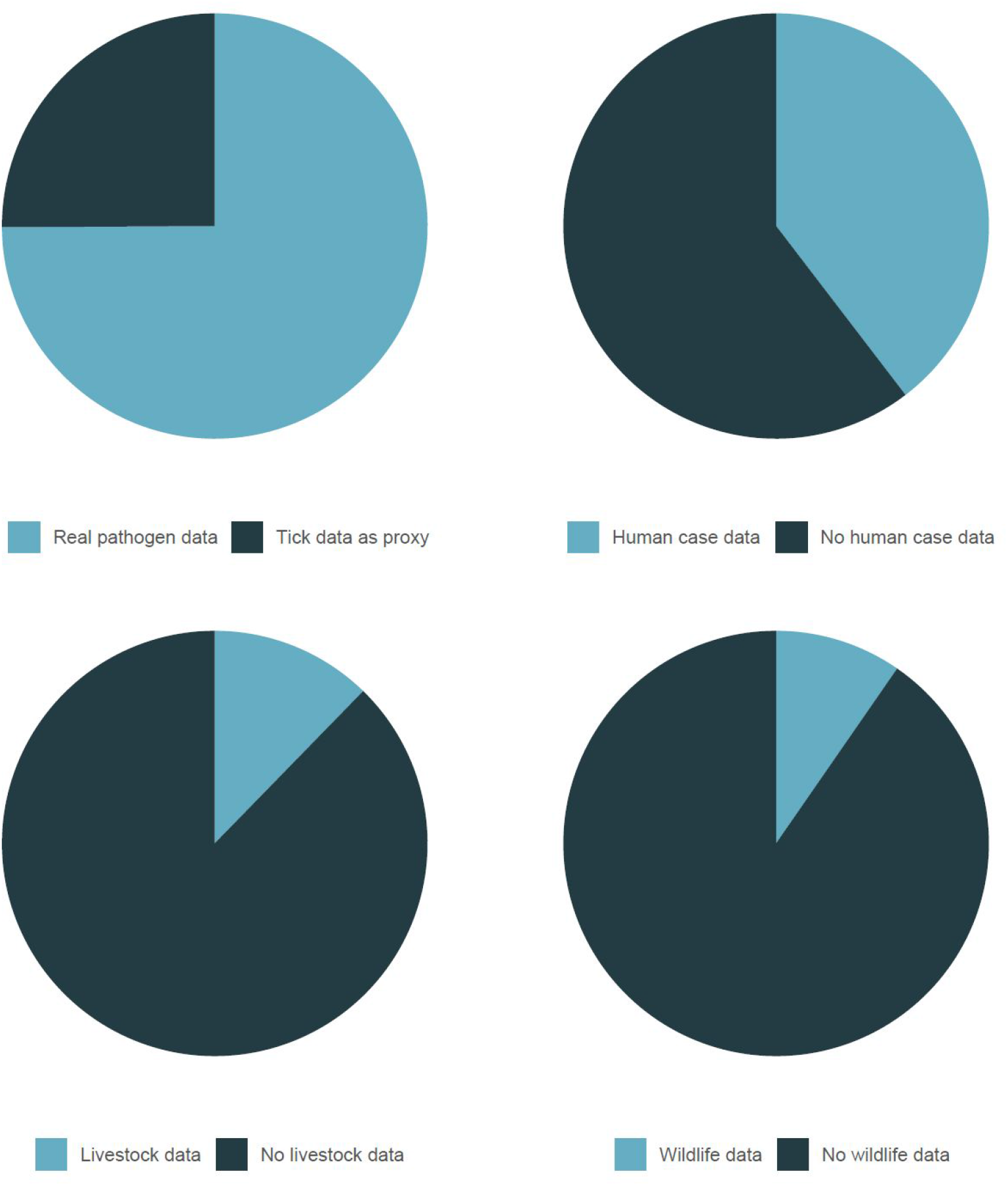
The proportion of studies using different data sources to generate maps of tick-borne disease distribution, transmission, or risk.

**Figure 5.**
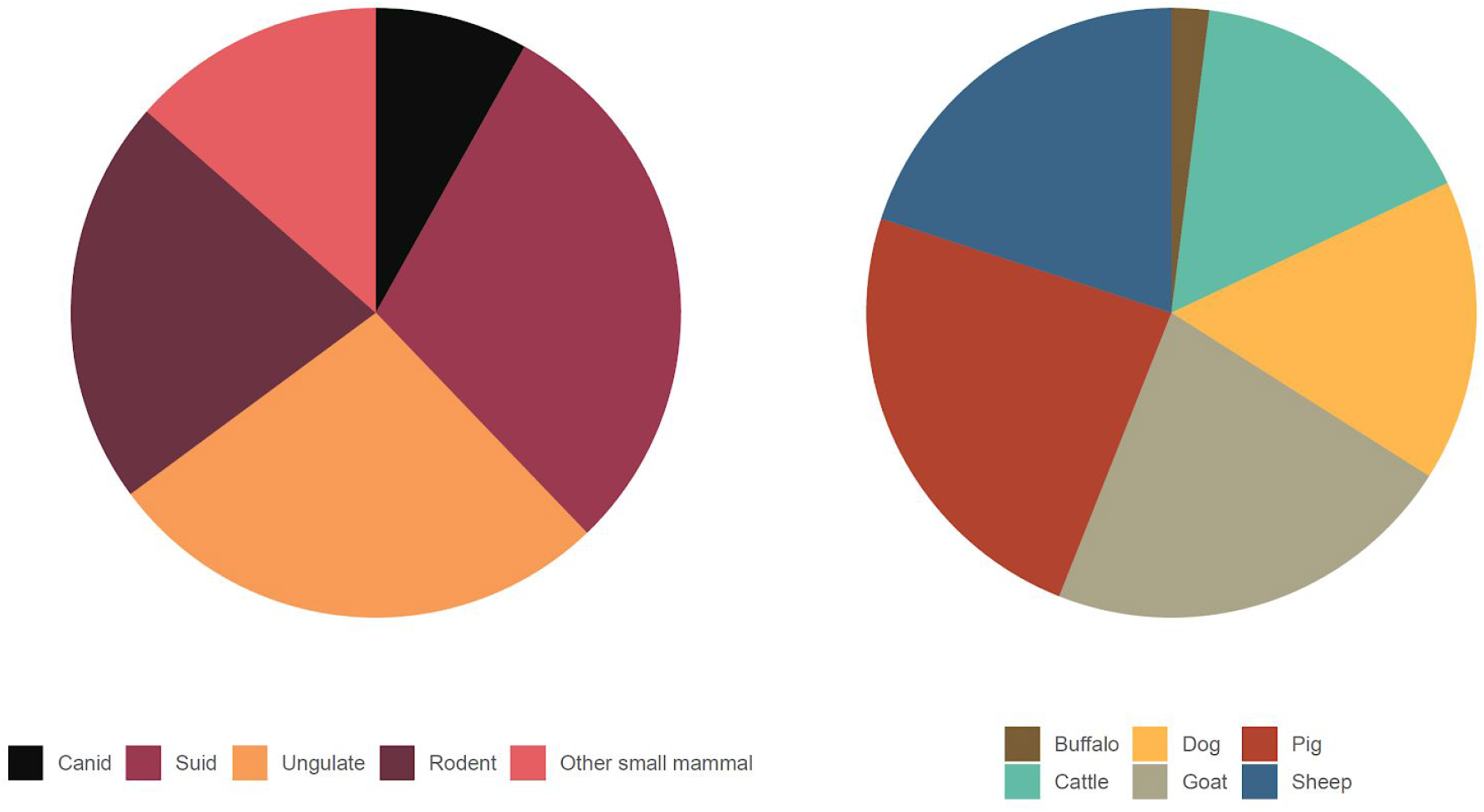
Proportion of studies with data on different wildlife (left) and livestock species (right).

The geographic foci and extent of studies included in the final dataset varied considerably, ranging from highly localized areas to mapped outputs with global extent. We found four global mapping studies on tick-borne pathogens, and a number of explicitly continental studies focused in Europe (n=20), Africa (n=4), the eastern Mediterranean (n=2), and Asia (n=1). Regionally, North American locations were heavily represented in the literature, where 35.31% of studies were conducted in the United States, and 11.55% in Canada (Fig. 6). While global mapping efforts are comparably low compared to North America, there are conspicuous regional gaps in mapped tick-borne disease studies, notably in portions of South America, Africa, the Middle East, central Asia, and southeast Asia.

**Figure 6.**
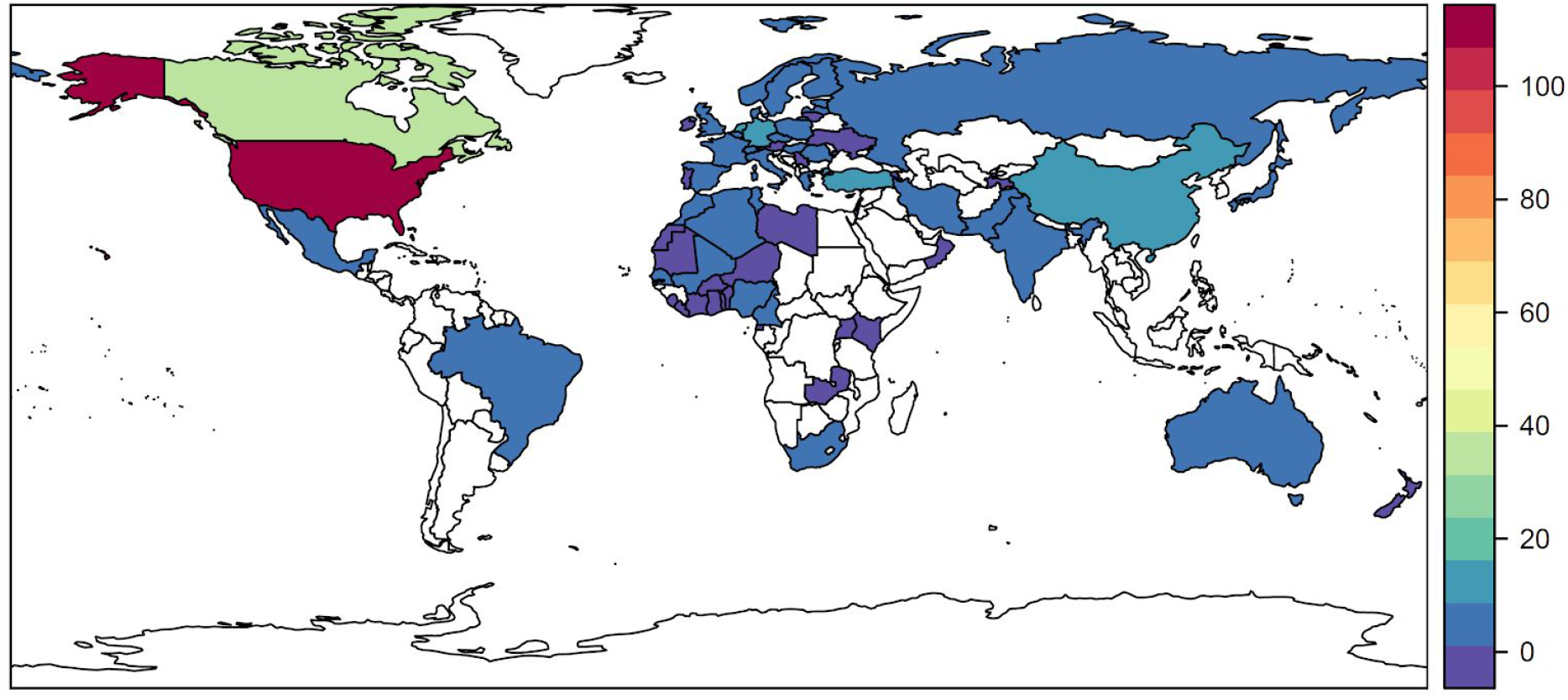
Number of studies describing the geography of tick-borne disease by country, excluding a handful of explicitly continental studies (most notably 20 in Europe, as well as four in Africa, two in the eastern Mediterranean, one in Asia, and four global mapping studies).

## Discussion

In this study, we performed a systematic review of scientific literature that has mapped tick-borne diseases, and quantified our findings in terms of distribution among pathogens, vectors, methods, geographic scope, and other attributes. Together, these provide a reasonable approximation of the current literature’s coverage of tick-borne diseases. Our review has demonstrated marked increases in both the number and diversity of work with spatial foci. Still, we have identified gaps in our geographic knowledge of tick-borne diseases. In many instances, basic natural history research to characterize pathogens and vectors will be important to improve the utility of risk mapping for understudied transmission systems. Efforts to expand surveillance of lesser-known pathogens, document sylvatic cycles, and increase the capacity for tick-borne disease surveillance in underrepresented regions will also help support future public health work.

### Why tick-borne diseases are difficult to map

Maps are commonly used to provide a tangible (and graphical) perspective on the “where” of disease risk, and can be used as part of the surveillance, prevention, and intervention toolbox in public health. Maps of vector-borne diseases carry an additional layer of complexity, as transmission risk is a combination of the abundance and behavior of vectors, the presence of the disease, and the opportunity for human infection. The data streams to describe vector and pathogen distributions often arise from data collection in historically disparate fields (i.e., entomology and infectious disease epidemiology respectively). In some instances, key vectors for zoonoses also remain unknown, as is the case for Crimean-Congo haemorrhagic fever virus (Okely et al. 2020), which may have different principal tick vectors by region. Resolving these data barriers and knowledge gaps is one step towards better geospatial studies.

Tick-borne diseases also pose a unique problem, given that all tick-borne viruses are zoonotic, which necessitates a view of their emergence and risk landscapes based on sylvatic cycles. For comparison, these transmission cycles are very well researched for some mosquito-borne diseases, such as yellow fever, and knowledge of pathogen-vector-host relationships can be used to improve risk mapping efforts (Jentes et al. 2011). While we have a firm understanding of sylvatic cycles for some tick-borne diseases, namely Lyme disease, that operationalized view of transmission does not exist for most tick-borne pathogens, and vertebrate hosts were only considered in a fraction of studies in our literature database. Data gaps are again characterized by disciplinary divides, as wildlife disease surveillance usually occurs separately from acarological collections, within-vector pathogen surveillance collection, and human public health records collections. Gaps in our knowledge regarding transmission cycles therefore present a major obstacle to quantifying and mapping risk of exposure.

### How tick-borne diseases are mapped

Although we recorded a tremendous diversity of approaches, we found that simple occurrence maps (i.e., displaying raw data points for either pathogens or their vectors) were the most common form of spatial data visualization. Dot maps of disease cases have long been used in epidemiology to communicate basic spatial information, and they remain a frequently used mapping approach that may complement more advanced quantitative methods (Smith et al. 2015). We also found that approximately one quarter of the studies in this review relied on tick presence as a proxy for pathogen presence and transmission risk. This is an intuitive way to formalize knowledge about the geographic range of risk when pathogen distributions are poorly sampled or unknown. However, maps derived solely from vector data underscore a clear need to refine perceptions of geographic risk through sampling efforts that focus on pathogens.

Mapping studies that employed more analytical approaches for spatial statistics or modeling (such as interpolated risk mapping, ecological niche models, and cluster analysis) have become more common especially in the past five years, likely due to advances in the diversity of modeling algorithms, availability of open source software, and increasing adoption of these methods in disease ecology. Perhaps most of all, we observed that these approaches relied on the existence of an ecosystem of open, accessible raw data describing the occurrence of ticks, pathogens, and clinical and veterinary cases. This secondary use of data was perhaps most evident in ecological niche modeling studies, where species presence data is commonly used as input for modeling algorithms (Elith and Leathwick 2009). This practice, while pragmatic, comes with the caveat that much work on establishing spatial risk of tick-borne diseases is hinged on a relatively small pool of existing data. This problem is exacerbated when diseases are rare events under current surveillance practices, or when tick vectors are challenging to sample, such as soft ticks in the genus *Ornithodoros* (Donaldson et al. 2016). We therefore recommend an emphasis on novel data collections, when possible, in future research.

Finally, we noted that despite substantial interest in the expansion of tick-borne diseases over time, there was fairly limited work that conclusively established this pattern. We encountered hundreds of examples of something akin to risk maps for tick-borne diseases, but most are so different in input and mapping technique as to be incomparable across studies, a lack of intercomparability that can stymie attempts to describe change over time. This can be addressed by direct work using a combination of modeling and endemicity mapping to update historical or baseline distributions, and project future areas of vulnerability; like other work, we note that this kind of work is heavily reliant on detailed, real-time primary data.

### Which tick-borne diseases are mapped

A small number of tick-borne diseases have been exceptionally well-studied and well-mapped. We found a preponderance of studies with information about Lyme disease and its vectors. Lyme disease has become the most frequently reported vector-borne disease in the United States, Canada, and Europe, a trend which underlies the geographic distribution of the research identified in this study (Lindgren and Jaenson 2006, Shapiro 2014, Lindsay 2016). The prevalence of Lyme disease mapping studies in the literature is unsurprising, as Lyme disease has been previously identified as a major research target, both in disease ecology and public health efforts (Han and Ostfeld 2019, Mac et al. 2019). Similarly, TBEV is also prioritized, likely due to its relative prevalence in humans, long history of its presence as a livestock related issue, and the intensity of research on this particular disease in Russia (National Academies of Sciences, Engineering, and Medicine et al. 2016, Ruzek et al. 2019, Bojkiewicz et al. 2020). The wealth of existing data for Lyme disease and TBEV, combined with ongoing surveillance efforts, translate into transmission systems that are extensively mapped across spatial and temporal scales, compared to other tick-borne diseases.

With these few exceptions, the majority of tick-borne pathogens are undermapped. Expanded pathogen diversity in mapping studies is mostly relegated to the past decade, a period which coincides with gains in the knowledge of tick-borne pathogen taxonomy, increased awareness of burden, and heightened public health interest (Vayssier-Taussat et al. 2013, Eisen and Eisen 2018, Pollet et al. 2020). Nevertheless, pathogens better represented in mapping studies are typically those that share common vectors with extensively studied pathogens. For example, pathogens that are also transmitted by *I. scapularis* and *I. ricinus* (the primary vectors of Lyme borreliosis and TBEV, respectively) tended to be better described in our data, often as part of integrative surveillance focused on these specific vectors instead of any one pathogen. Pathogens capable of transmission through agricultural production systems were also the focus of many mapping studies, even when the human burden of zoonotic transmission is comparatively low, as is the case with African swine fever and Q fever. In these instances, the bulk of mapped studies stem from the existence of established surveillance in livestock, management of wildlife populations, or testing of agricultural products (e.g. bulk tank milk testing) (Hilbert et al. 2015, Food and Agriculture Organization of the United Nations 2019).

In this review, we find several pathogens of increasing public health importance that would make good candidates for targeted surveillance efforts, where areas of transmission risk are largely derived from vector distributions. For example, four pathogens - Omsk hemorrhagic fever virus, sawgrass virus, Colorado tick fever virus, and *Rickettsia africae* - were entirely absent from our search, and represent important future priorities. Similarly, understudied pathogens transmitted by *I. scapularis* - including *A. phagocytophilum, E. chaffeensis*, and Powassan virus - could be more regularly involved in vector surveillance efforts. In a similar vein, the geographic distribution of reviewed literature indicates several regional disparities in mapped tick-borne disease research, highlighting potential opportunities for increased research efforts. South America, for example, is poorly represented in the tick mapping literature, despite having confirmed cases of tick-borne diseases and known tick species of medical importance (Guglielmone et al. 2006). Documentation of zoonotic pathogens on the continent is similarly underfunded, and tick-borne transmission cycles are an active area of research (Guglielmone et al. 2006, Rodriguez-Morales et al. 2018). Identifying regional priorities for surveillance based on clinical and veterinary significance, and expanding the purview of tick-borne disease mapping using participatory approaches alongside quantitative and GIS work, will help manage the burden of tick-borne disease where it remains the highest.

## Data Availability

All data and code are available at github.com/viralemergence/tickmaps

## Author Contributions

All authors contributed to the conception, design, and writing of the study. CAL and ALW gathered data, and CJC generated data visualizations.

## Acknowledgements

CJC and SJR were supported by funding to the Viral Emergence Research Initiative (VERENA) consortium including NSF BII 2021909. CAL, HDG, and SJR were funded by NIH 1R01AI136035-01. ALW and SJR were additionally funded by CDC grant 1U01CK000510-01: Southeastern Regional Center of Excellence in Vector-Borne Diseases: The Gateway Program. This publication was supported by the Cooperative Agreement Number above from the Centers for Disease Control and Prevention. Its contents are solely the responsibility of the authors and do not necessarily represent the official views of the Centers for Disease Control and Prevention.

## Data and Code Availability

All data and code are available at github.com/viralemergence/tickmaps

## Notes

### Competing Interest Statement

The authors have declared no competing interest.

